# High-throughput quantitation of SARS-CoV-2 antibodies in a single-dilution homogeneous assay

**DOI:** 10.1101/2020.09.16.20195446

**Authors:** Markus H. Kainulainen, Eric Bergeron, Payel Chatterjee, Asheley P. Chapman, Joo Lee, Asiya Chida, Xiaoling Tang, Rebekah E. Wharton, Kristina B. Mercer, Marla Petway, Harley M. Jenks, Timothy D. Flietstra, Amy J. Schuh, Panayampalli S. Satheshkumar, Jasmine M. Chaitram, S. Michele Owen, M. G. Finn, Jason M. Goldstein, Joel M. Montgomery, Christina F. Spiropoulou

**Affiliations:** Viral Special Pathogens Branch, Division of High-Consequence Pathogens and Pathology, Centers for Disease Control and Prevention, 1600 Clifton Rd, Atlanta, GA 30329, USA; School of Chemistry and Biochemistry, School of Biological Sciences, Georgia Institute of Technology, 901 Atlantic Dr., Atlanta, GA 30332, USA; Reagent and Diagnostic Services Branch, Division of Scientific Resources, Centers for Disease Control and Prevention, 1600 Clifton Rd, Atlanta, GA 30329, USA; Emergency Response Branch, Division of Laboratory Sciences, Centers for Disease Control and Prevention, 4770 Buford Hwy., Atlanta, GA 30341, USA; Newborn Screening & Molecular Biology Branch, Division of Laboratory Sciences, Centers for Disease Control and Prevention, 4770 Buford Hwy., Atlanta, GA 30341, USA; Poxvirus and Rabies Branch, Division of High-Consequence Pathogens and Pathology, Centers for Disease Control and Prevention, 1600 Clifton Rd, Atlanta, GA 30329, USA; Division of Laboratory Systems, Centers for Disease Control and Prevention, 1600 Clifton Rd, Atlanta, GA 30329, USA; National Center for HIV/AIDS, Viral Hepatitis, STD, and TB Prevention, Centers for Disease Control and Prevention, 1600 Clifton Rd, Atlanta, GA 30329, USA

## Abstract

SARS-CoV-2 emerged in late 2019 and has since spread around the world, causing a pandemic of the respiratory disease COVID-19. Detecting antibodies against the virus is an essential tool for tracking infections and developing vaccines. Such tests, primarily utilizing the enzyme-linked immunosorbent assay (ELISA) principle, can be either qualitative (reporting positive/negative results) or quantitative (reporting a value representing the quantity of specific antibodies). Quantitation is vital for determining stability or decline of antibody titers in convalescence, efficacy of different vaccination regimens, and detection of asymptomatic infections. Quantitation typically requires two-step ELISA testing, in which samples are first screened in a qualitative assay and positive samples are subsequently analyzed as a dilution series. To overcome the throughput limitations of this approach, we developed a simpler and faster system that is highly automatable and achieves quantitation in a single-dilution screening format with sensitivity and specificity comparable to those of ELISA.

**One sentence summary:** Protein complementation enables mix-and-read SARS-CoV-2 serology that rivals sensitivity and specificity of ELISA but excels in throughput and quantitation.

## Introduction

Detecting viral nucleic acids, primarily by reverse-transcription quantitative polymerase chain reaction (RT-qPCR), is the method of choice for diagnosing acute viral infections. However, diagnosing acute infections does not account for asymptomatic infections or bias resulting from availability of care or individuals not seeking care *(1)*. Therefore, serology plays an important part in completing the epidemiological picture when a new pathogen, such as severe acute respiratory syndrome coronavirus 2 (SARS-CoV-2) *(2, 3)*, emerges. Analyzing humoral responses in convenience samples is a quick way to estimate how many individuals have already been exposed to a pathogen. On the other hand, designing community-level and special population studies offers the opportunity to solicit tailored participant information, such as symptom history, for correlation analyses *(1)*. Apart from surveys that aim to produce generalizable information, serology has also been used to link together individual COVID-19 clusters where nucleic acid evidence was absent *(4)*.

While the serological studies mentioned above could potentially be conducted with qualitative serology assays (which report presence or absence of SARS-CoV-2 antibodies), answering other pertinent questions requires quantitative determination of antibody levels. One timely question is the possibility of waning immunity to SARS-CoV-2 and reinfection with this pathogen. Some studies have called into question the long-term stability of the humoral immune response against this virus *(5–7)*, and small-scale human challenge studies with a benign human coronavirus have shown that at least asymptomatic reinfection with the same strain *(8)* or a related one *(9)* is possible as soon as a year after initial infection. Meanwhile, other studies did not observe rapid waning of the humoral response *(10, 11)* and yet others have evaluated the immune signature of mild SARS-CoV-2 infections and predicted that this immune memory is likely to be protective *(12)*. Longitudinal sampling combined with accurate serological quantitation is necessary to monitor variations in antibody levels associated with waning immunity or reinfection *(13)*. Vaccine trials also require quantitative serology to establish immune correlates and to decide on optimal vaccine dose and regimen.

Another timely question is the role that other animal hosts may play in the future of the SARS-CoV-2 pandemic. To date, the natural reservoir or intermediate hosts leading to the emergence of this virus in humans have not been definitively identified. However, cats *(14)*, deer mice *(15)*, ferrets, fruit bats *(16)*, and golden hamsters *(17, 18)* can transmit the virus in a laboratory setting. Furthermore, SARS-CoV-2 outbreaks have occurred in farmed mink, and virus phylogenetic evidence suggest that mink may have transmitted the virus back to humans *(19)*. One-health investigations into the reservoir species and potential intermediate hosts of SARS-CoV-2, as well as into spillover events into species with which people have frequent contact, would benefit from species-independent serology methods to determine the role these species play in virus transmission and spread in human populations.

Protein complementation (also referred to as protein fragment complementation) is an assay principle primarily used for studying protein-protein interactions in cells. The interacting proteins are coupled to protein or peptide moieties that generate a detectable signal only when brought into close proximity with each other. Fluorescence-based complementation assays can provide information on protein-protein interactions and on the localization of the interaction in living cells. Such assays have been developed by dividing the structure of the green fluorescent protein and its relatives into fragments that only fluoresce when in close proximity. Luciferase-based systems, however, are generally more sensitive than fluorescent protein systems *(20, 21)*. The recent such splitting of NanoLuc, the most sensitive luciferase to date *(22)*, enabled us to build a serological assay that matches the sensitivity and specificity characteristics of classical enzyme-linked immunosorbent assays (ELISAs) but also offers easier automation, higher throughput, and independence of host species while quantitating antibodies in a single step.

## Results

### Assay principle

We hypothesized that antibodies, having two or more antigen binding sites (depending on class), could act as ‘protein complementation bridges’ to bring together antigens in solution, resulting in a detectable signal (Fig. 1A). The receptor-binding domain (RBD) of SARS-CoV-2 spike protein was chosen as the assay antigen, since it is known to offer high specificity against prevalent antibodies against common human coronaviruses *(23, 24)*. Two RBD-fusion proteins, each with one fragment of the split NanoBit luciferase *(22)*, were produced in mammalian cells and purified. To test if binding of both fragments simultaneously could indeed result in protein complementation, we compared the signal generated by a monoclonal antibody (mAb) either in its intact form, or as F(ab′)_2_ or Fab fragments produced by protease digestion. Signal could be observed when intact mAb and F(ab′)_2_ were used, but not with Fab fragments, indicating that antibody binding two antigens simultaneously is necessary for assay activation (Fig. 1B). All three forms of the mAb retained the ability to inhibit interaction of RBD with its receptor, ACE2 (angiotensin-converting enzyme 2), showing that despite being unable to activate the assay the Fab fragment retained binding affinity for the RBD antigen (Supplementary Fig. 1A and B).

**Fig. 1.**
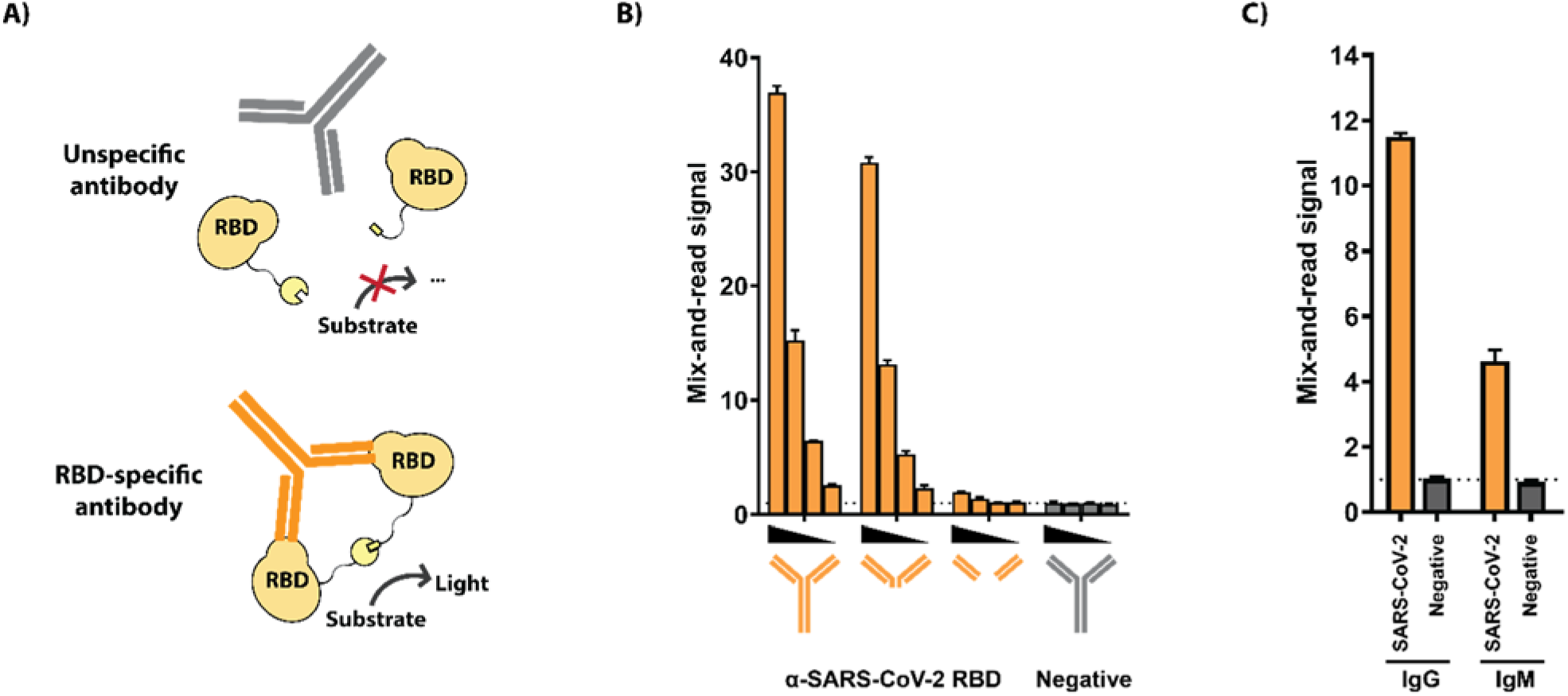
Assay principle. A) Schematic. Small and large fragments of the NanoBit split luciferase are tethered to SARS-CoV-2 receptor-binding domains (RBD) via peptide linkers. When a specific antibody binds 2 RBD antigens, it forces the luciferase halves into proximity with each other, thereby activating the enzyme. Signal is quantified from a homogeneous assay setup after adding luciferase substrate. B) Demonstration of assay principle. Dose-responsive assay activation by an RBD-specific monoclonal antibody or corresponding F(ab′)_2_ or Fab fragments (orange) or control antibody (gray). C) Activation by specific IgG and IgM antibodies. Human IgG and IgM fractions were purified from a SARS-CoV-2 specific serum pool or a negative control pool and tested for activity in the assay. B and C: technical triplicates with averages and standard deviations presented.

Since signal generation is dependent on an antibody binding two RBD antigens but does not require the Fc region, we next investigated whether purified antibodies from convalescent COVID-19 patients yield a signal and whether the pentameric structure of IgM antibodies could also activate the RBD reporter pair. Total IgG and IgM fractions were purified from a convalescent COVID-19 serum pool and a control pool (Supplementary Fig. 2). The system could be activated by the specific IgG fraction and the specific IgM fraction, indicating that the assay measures total immunoglobulin reactivity against the RBD (Fig. 1C).

### Training set and cut-off determination

Assay performance was optimized by varying factors, including antigen concentrations and ratios, buffer and blocker choices, plastic materials, incubation times, and substrates. Additionally, we observed that using RBD domain consisting of SARS-CoV-2 residues 319–591 *(25)* offered lower background luminescence signal, and therefore higher sensitivity, than using residues 319–541 *(26)* (optimization data not shown).

In order to establish the assay in human diagnostics, we opted to use the two-step strategy, in which the assay cut-off was first established with a training sample set, and performance was then validated with an independent sample set. The training set consisted of 288 serum samples: 85 samples from individuals with prior SARS-CoV-2 infection confirmed by nucleic acid testing, and 203 negative samples (117 negative control samples sourced for the purpose, 100 of which were collected prior to SARS-CoV-2 emergence, and 86 residual serum samples collected in the U.S. between 2016 and 2019 for hantavirus diagnostics). These training set samples were tested with the optimized mix- and-read assay and with ELISA against RBD with the same antigen length (spike residues 319–591). The mix-and-read assay signals obtained from single serum dilution varied by 3 orders of magnitude (Fig. 2A). The values were categorized for receiver operating characteristics (ROC) analysis based on presence or absence of prior PCR-confirmed infection (Fig. 2B and 2C). We decided to prioritize assay specificity and chose the cut-off value of 2.5, which enabled the highest sensitivity (antibodies detected in 92.9% of the samples with an earlier SARS-CoV-2 nucleic acid finding) while maintaining 100% specificity. We noticed that the average negative result was somewhat below 1, indicating modest signal inhibition by serum. The high cut-off value (>6 standard deviations above the average of negative sample signal) also minimizes impact of user error in case sample is omitted. Agreement between ELISA and the mix-and-read assay was very high (kappa value 0.949; 95% confidence interval 0.908– 0.989). The mix-and-read assay found no antibodies in 6 of 85 samples with confirmed SARS-CoV-2 infection; 4 of these samples were also negative by RBD ELISA; 1 was ELISA positive with a weak signal and 1 was ELISA positive with a moderate signal.

**Fig. 2.**
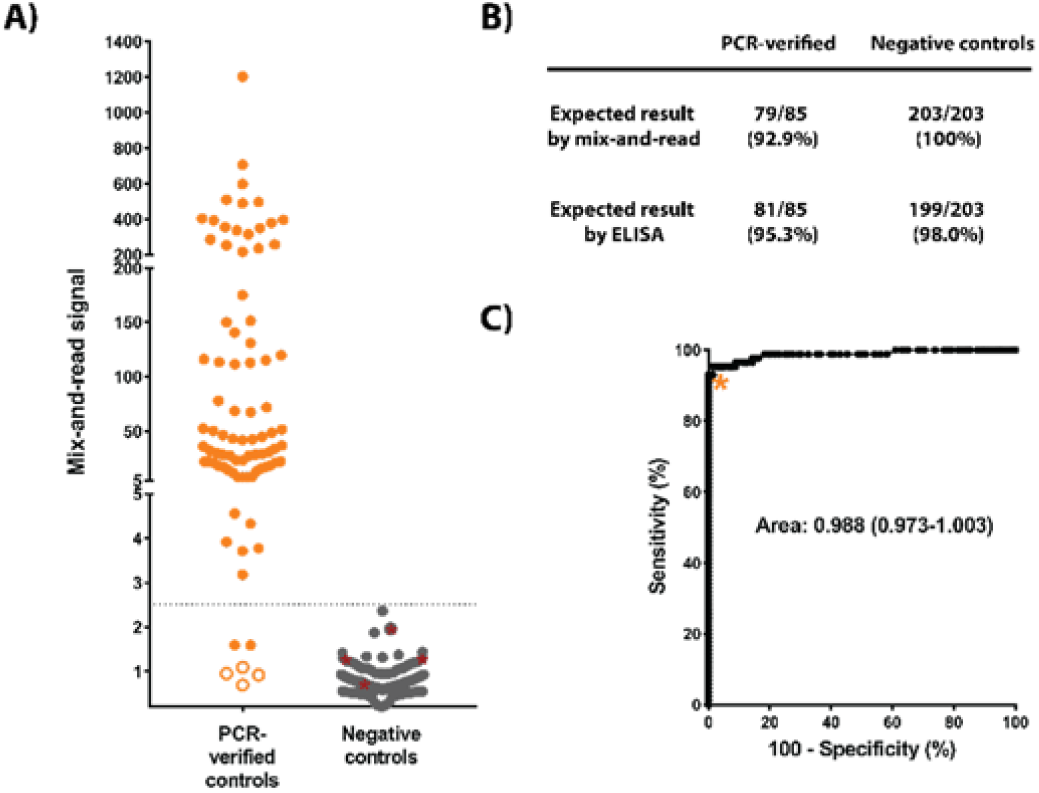
A) Training set data. Serum samples from cases with earlier nucleic acid confirmed SARS-CoV-2 infection and samples considered true negatives (no COVID-19 diagnosis, or sample collected prior to SARS-CoV-2 emergence) were analyzed by the mix-and-read assay and by RBD ELISA. Out of 85 positive controls, 79 were above proposed cut-offs on both ELISA and mix-and-read assays (orange symbols above signal 2.5), 4 gave no signal distinguishable from negative controls on either assay (open orange symbols), and 2 were positive on ELISA only (orange symbol below 2.5). All negative control signals (N = 203) were below proposed mix-and-read cut-off (gray symbols), but 4 of those samples were positive by ELISA (gray symbols highlighted by red asterisks). B) Summary of the training set data. C) Receiver operating characteristics (ROC) analysis of the data and cut-off selection. Cut-off value of 2.5 (as fold over blank samples) was chosen by ROC analysis of the training set data so that 100% specificity could be maintained.

While serum is the standard sample matrix for serology, we wanted to establish whether plasma could be tested as well. Paired human serum and EDTA plasma samples gave similar signals in the mix-and-read assay, suggesting that EDTA plasma samples could also be analyzed (Supplementary Fig. 3A). Furthermore, testing mouse sera from animals immunized with SARS-CoV-2 spike antigens formally showed that the assay works with non-human serum as well (Supplementary Fig. 3B).

### Independent validation set

Using the cut-off value established with the training set, we tested 101 more serum samples from individuals with previous confirmed SARS-CoV-2 infection and 97 samples from negative control individuals (96 of which were collected before the outbreak) (Fig. 3A). The RBD ELISA gave a positive result for 91 of 101 positive control samples, while the mix-and-read results were positive for 86 of those samples. Conversely, the mix-and-read assay correctly identified the 97 negative samples, maintaining 100% specificity, while the RBD ELISA gave negative results for 95 of 97 negative controls (97.9% specificity) (Fig. 3B). The independent set results from the two assays were in high agreement (kappa value 0.929; 95% confidence interval 0.877–0.980). Repeat mix-and-read analysis of 100 samples exhibited high day-to-day signal reproducibility (Fig. 3C), as did 3 positive quantitation controls and a negative control that were used to monitor run-to-run reproducibility (Supplementary Fig. 4). Combined ELISA results and ROC analysis are presented in Supplementary Fig. 5.

**Fig. 3.**
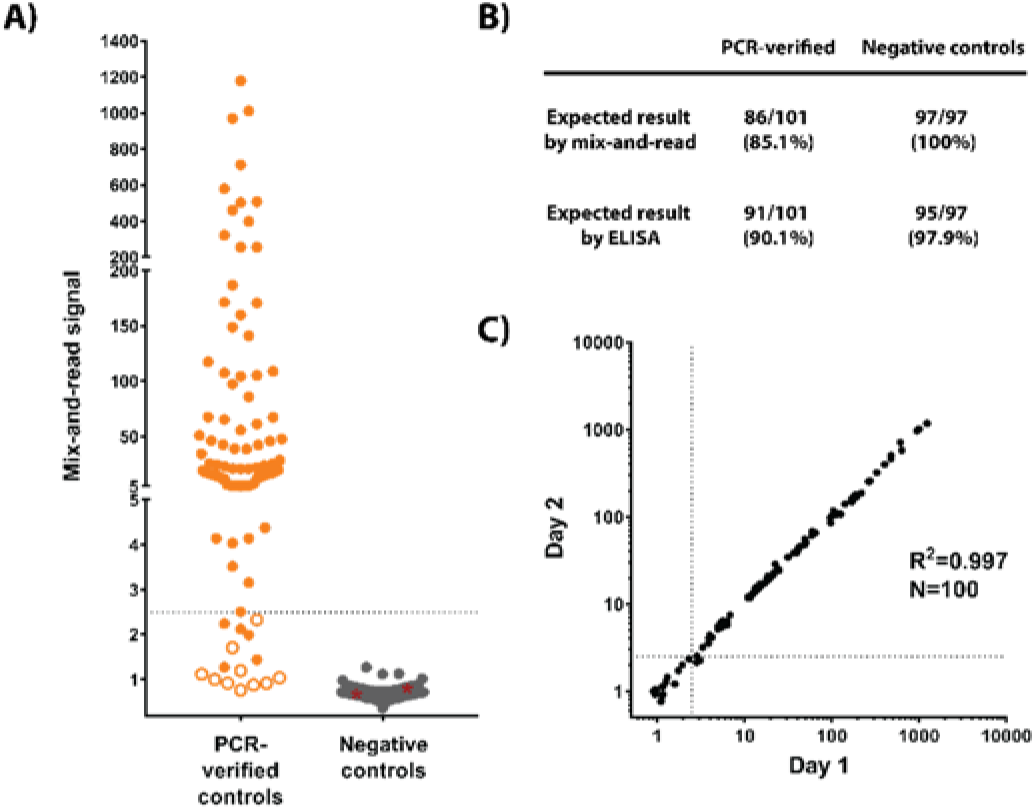
Independent validation set data. A) Serum samples from confirmed COVID-19 cases and samples considered true negatives (collected in 2020 but without SARS-CoV-2 detection by nucleic acid testing, N = 1; or collected prior to COVID-19 outbreak, N = 96) were analyzed by RBD ELISA and by the mix-and-read assay using cut-off values determined with the training set. Out of 101 samples with positive SARS-CoV-2 history, 86 were found positive by the mix-and-read assay (orange symbols above signal 2.5), 10 were negative on both assays (open orange symbols), and 5 were positive on ELISA only (orange symbol below 2.5). All negative controls (N = 97) were negative on the mix-and-read assay (gray symbols), while 2 of those samples were positive on ELISA (gray symbols highlighted by red asterisks). B) Summary of the independent set results. C) Day-to-day reproducibility of the mix- and-read assay. Samples from the independent validation set were tested on 2 consecutive days and correlation analysis performed on log-transformed values. Samples that gave a discordant qualitative result between the days (near the cut-off, above one day and below the other) were judged negative for purposes of sensitivity and specificity calculations.

### Linear range

The assay principle used sets a theoretical upper limit to antibody quantitation. If specific antibodies (or, more precisely, high-affinity antigen binding sites) exceed RBDs, then the number of antibody molecules binding 2 RBDs will decline whereas the number of antibodies binding only 1 or no RBDs will increase. Therefore, increasing the antibody concentration excessively may actually result in declining signal. To test the theory in practice, monoclonal antibodies were spiked into non-reactive human serum at concentrations between 1 ng/mL and 100 µg/mL. Dose response curves showed that maximal signal was obtained at mAb concentrations 10–30 µg/mL, corresponding to 1:1 to 1:3 stoichiometry between the RBDs and the antigen binding sites (Fig. 4A). Increasing the mAb concentration to 100 µg/mL resulted in lower, though strong signal, suggesting that correct qualitative results would be obtained with even the most reactive clinical samples.

**Fig. 4.**
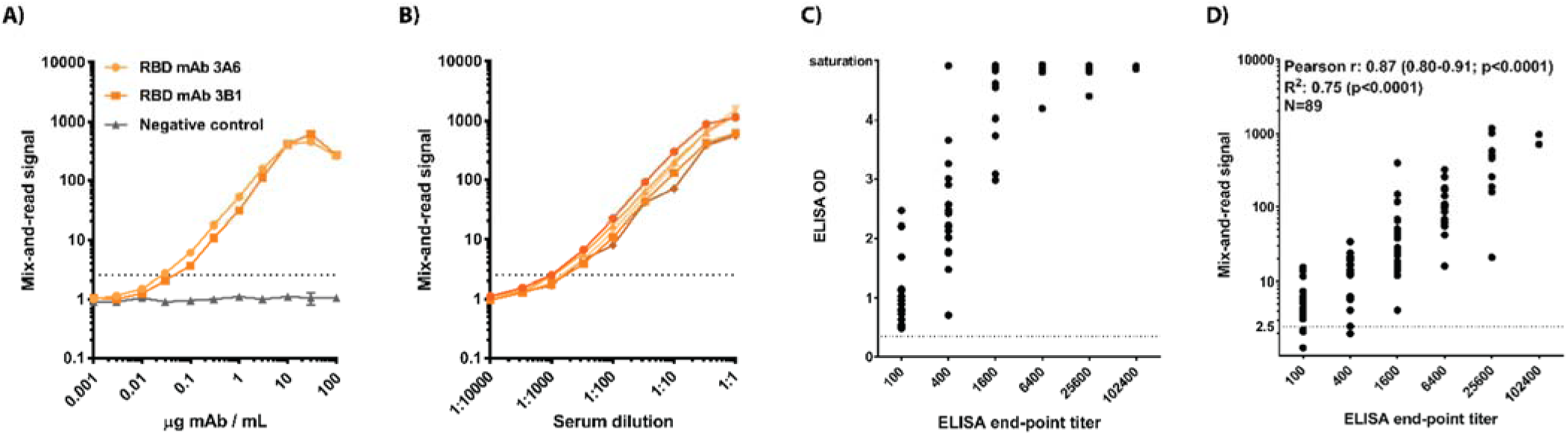
Quantitation. A) Two positive control IgG mAbs and a negative control mAb were titrated to determine the linear range of the mix-and-read signal. B) Five serum samples that gave a strong signal in initial testing were titrated in negative human serum to determine if they reached the saturation level identified by mAbs. C) Correlation of ELISA end-point titer and ELISA optical density from 1:100 sample dilution. D) Correlation of ELISA end-point titer and mix-and-read signal. Correlation analysis (Pearson r and 95% confidence interval) and linear regression were performed on log-transformed mix-and-read data and log2-transformed ELISA end-point titers. A and B: technical duplicates with averages and ranges presented.

### Single-dilution quantitation with the mix-and-read assay

Titration results with monoclonal antibodies suggested that the assay signal saturates and declines when excessive antibody concentrations are used. To test if the same phenomenon could impact SARS-CoV-2 RBD antibody quantitation in clinical serum samples, 5 samples with high signal-to-background ratios were serially diluted in non-reactive human serum and dose-response curves were generated. All samples gave the strongest signal when undiluted, showing that even the most reactive samples among the >180 positives identified here do not contain high enough amounts of specific antibodies to saturate the assay. However, the shape of the dose-response curves near 1:1 dilution suggest that these strong samples are approaching the higher quantitation limit (Fig. 4B).

To investigate if SARS-CoV-2 RBD antibody titers could be inferred from a single screening dilution, end-point ELISA titers were determined for 89 positive samples of the validation set. Plotting ELISA optical density (OD) values from the screening dilution of 1:100 against the end-point titer showed the expected correlation at low dilutions but loss of quantitation in samples that reached saturation OD in the screening (Fig. 4C). In contrast, no apparent saturation was seen in the mix-and-read assay, and those values and ELISA end-point titers correlated at Pearson r 0.87 (95% confidence interval 0.80–0.91, p < 0.0001) and R^2^ 0.75 (p < 0.0001), indicating that sample screening at a single dilution is predictive of ELISA endpoint titer in a semi-quantitative manner (Fig. 4D).

## Discussion

With nearly 500 samples analyzed in parallel, our mix-and-read assay results and RBD ELISA results were in excellent agreement. The observed specificity and sensitivity characteristics are comparable to those of independently evaluated, high-performing commercial assays *(27, 28)*, although formal comparisons are best done with standardized validation sample sets.

The mix-and-read assay identified RBD antibodies in 88.7% of the samples with history of confirmed SARS-CoV-2 infection, while our RBD ELISA identified antibodies in 92.5% of those samples. The time interval from symptom onset to sampling was known for 9 of the 14 samples that were found seronegative by both assays and ranged from 14 to 30 days, suggesting that time of sampling was not the cause of negative results. Since the mix-and-read assay can detect IgM antibodies as well as IgG antibodies, class switching should not be required for detecting seroconversion. However, in general, the lower physiological concentration of IgM antibodies may render them the less potent assay activator as compared to IgG antibodies. In conclusion, the implied seroconversion rate of ∼90% is comparable to rates reported by others *(5, 10, 29)*, although universal seroconversion has also been reported *(30)*.

The mix-and-read assay showed 100% specificity in this sample set while the specificity of ELISA was 98%. The wide dynamic range of the mix-and-read assay allowed us to keep the cut-off value over 6 standard deviations above the mean of the negative controls. Although apparent sensitivity could be marginally improved and the correlation of the two assays further increased by lowering the threshold and accepting false positives at low frequency, we chose to use a cut-off that emphasizes specificity. It is important to remember that, especially in populations in which antibody prevalence is low, the positive predictive value (likelihood that a positive result is correct) deteriorates rapidly when specificity falls below 100% *(31, 32)*. For example, a test with 99% specificity and high sensitivity may be useful for many purposes in an area of high prevalence, but in an area where true prevalence is 1%, half of the positive test results would be false. Of note, 2 of the 6 negative control samples that produced an ELISA signal were near the chosen ELISA cut-off, but 4 (collected prior to recognized SARS-CoV-2 emergence) gave moderate to strong OD values, implying cross-reactivity between the RBD antigen and unknown antigens. Interestingly, despite the high overall correlation between the two assays, these ELISA-reactive samples were negative when read on the mix-and-read assay (3 with signal < 1, one being close to the cut-off value 2.5). The reason for this phenomenon is unknown, but increased specificity may arise from factors like the strict requirement for simultaneous binding of 2 antigens, better preservation of antigen structure in solution than on plate surface, or steric masking of cross-reactive epitopes by the split luciferase moieties.

One limitation of our study is that the work was conducted primarily to evaluate a new diagnostic approach. Control samples were sourced based on availability and not as part of a serological survey, and some associated data, including demographics, accurate location, symptoms, underlying conditions, and hospitalization history of the individuals were absent or incomplete and are not reported here. Furthermore, we note that the samples originated in the U.S., and would encourage testing representative control cohorts any time a serological test is implemented in a new area or population, or indeed species, to account for potential differences in baseline reactivity. Of note, we found that convalescent SARS-CoV-1 sera can also activate the assay (data not shown). While this has little practical relevance in human diagnostics due to the small number of convalescent SARS-CoV-1 patients, it is noteworthy with regards to ecological investigations and instances where infection with a closely related virus is a possibility.

In summary, we used SARS-CoV-2 as an example to demonstrate that modern protein complementation enables design of simple yet robust serological assays that are easy to automate. Complementation detects antibodies due to their binding of multiple antigens simultaneously, and thus the assay has no apparent antibody class or species restrictions. The improved dynamic range means that more information can be derived from primary screening than is possible with ELISA. Complementation-based serology has the power to increase the output of human SARS-CoV-2 serology efforts and to advance One Health investigations.

## Materials and Methods

While this manuscript was in preparation, a qualitative commercial product utilizing the same principle was released (Promega). The two systems have been developed independently and the technical details of the commercial product are unavailable.

### Protein expression

Promega NanoBit split luciferase fragments LgBit (large bit) and SmBit (small bit) were fused to the N-terminus of SARS-CoV-2 isolate Wuhan-Hu-1 (GenBank MN908947) RBD. The sequences were separated by a flexible region consisting of a 5-residue linker (GGGGS), 8 × His, and a 16-residue linker (SGSSGGGGSGGGGSSG). IL-6 signal peptide was used to enable secretion. Different RBD lengths, including residues 319–541 and 319–591 of the viral glycoprotein spike were used in the optimization phase. All data presented here are from the 319–591 version of the RBD.

The DNA sequences for the above-described fusion proteins were cloned into an episomal vector (System Biosciences) into which a puromycin resistance cassette had been added to enable selection of pools stably expressing these proteins. To express the proteins, human Expi293 cells (Thermo Scientific), growing in Expi293 Expression medium, were transfected with FectoPRO reagent (PolyPlus Transfection). The supernatants were clarified by slow-speed centrifugation and filtered over polyethersulfone (0.2 micron) membranes, and His-tagged proteins were purified by immobilized metal affinity chromatography using HisTrap Excel nickel columns (Cytiva). After dialysis in PBS, the proteins were flash frozen in liquid nitrogen and stored at -80°C.

The proteins for RBD/ACE2 binding assay were produced the same way as above. Human codon-optimized ACE2 (residues 1-615, NM_001371415) was expressed with C-terminal LgBit using the same linker and His-tag sequence as described earlier. Similarly, RBD (residues 1-14 and 319-541) was expressed with C-terminal SmBit.

The RBD that was used as ELISA antigen consisted of residues 319-591 and was expressed with an N-terminal IL-6 signal peptide, 8 × His and 3C protease cleavage site. The protein was produced and His-purified as above, and the epitope tag removed by digestion with 3C protease. The resulting product was further purified by size-exclusion chromatography using Superdex 200 columns.

### Mix-and-read assay procedure

After initial optimization in a 96-well format, the assay was moved to 384-well plates, and all data presented here were produced using that setup. 20 ng of SmBit-SARS-2-RBD, and equivalent amount of LgBit-SARS-2-RBD in 1:1 molar ratio were added per well of 384-well plates in 20 µL 1% BSA/PBS (w/V) diluent. 10 µL sample was then added, and the mix was incubated for 1 h at room temperature. 30 µL Promega NanoGlo assay reagent was then added and the luminescence resulting from SmBit and LgBit proximity was quantified 10 min later using a Synergy Neo2 instrument (BioTek) with 0.5 s integration time and with gain set to 200 and read height to 7.5 mm. Wells with diluent instead of sample were used to determine the assay baseline. Samples with strong signal (up to 2–3 orders of magnitude above baseline) were removed and the plate was read immediately again to minimize error from signal leak between wells. Assay performance was monitored using 4 standards: normal human serum (from plasma) as a negative control, and RBD-specific mouse monoclonal antibody 3A2 spiked into the same serum at 3 different concentrations (10, 2.5, and 0.5 µg/mL).

### ELISA

ELISAs were conducted against in-house produced SARS-CoV-2 RBD antigen (spike residues 319–591). 50 ng of antigen was bound to Immulon 2 HB plates by overnight incubation in 100 µL PBS at 4°C. Wells without antigen were similarly prepared. Wells were washed 2× with 300 µL 0.1% Tween-20 in PBS (PBS-T) and blocked with 300 µL 5% (w/V) non-fat dry milk (CellSignaling Technologies) in PBS-T for 1 h at room temperature (RT). After 3 washes, samples diluted 1:100 in 100 µL 5% milk in PBS-T were added and incubated for 1 h at RT. The plates were washed 3×, and mouse secondary anti-human IgG antibody conjugated to HRP (Accurate Chemical #JMH035098) was added at 1:10.000 dilution in 100 µL 5% milk in PBS-T and incubated for 30 min at RT. After 3 washes, 100 µL TMB Ultra ELISA substrate (Thermo Fisher Scientific) was added and reactions were stopped after 20 min at RT with addition of 100 µL 1 M hydrochloric acid. Optical density was read at 450 nm and densities from antigen-containing wells were corrected by reducing the values obtained from respective no-antigen wells.

### RBD-ACE2 binding assay procedure

3.2 ng RBD-Smbit (residues 319-541, C-terminal tag) was diluted in 20 µL 0.02% BSA/PBS and mixed with 20 µL mAb dilution (17 µg/mL) or mAb fragment dilution (corresponding molarity) in 96-wells. After 1 h incubation at RT, ACE2-LgBit was added in volume of 20 µL so that 10× molar excess over the RBD was achieved. After 1 h incubation at RT, 60 µL NanoGlo reagent (Promega) was added and luminescence quantified as with the mix-and-read assay. Wells where antibody dilution was substituted with the diluent served to determine the baseline signal of RBD-ACE2 binding.

### Monoclonal antibodies and mouse sera

The monoclonal antibodies were generated by repeated mouse immunizations and cloned using standard methods. Polyclonal mouse serum from such immunizations was used to test the assay performance. Thorough description of these antibodies will be presented elsewhere (Chapman et al., in preparation). All animal procedures were approved by the Institutional Animal Care and Use Committee of the Georgia Institute of Technology.

Monoclonal antibody 3A2 was cleaved into F(ab′)_2_ and Fab fragments using Pierce Mouse IgG1 Fab and F(ab′)2 Preparation Kit (Thermo Fisher Scientific) according to manufacturer’s instructions. Briefly, the two fragments were generated by controlling the specificity of immobilized ficin by cysteine-HCl concentration and incubation time. Immobilized protein A was used to remove uncleaved mAb and Fc fragments. The fragments were concentrated with 500× buffer exchange against PBS using spin columns with 10 kDa molecular weight cut-off (Amicon, Millipore Sigma). mAbs were diluted to 10/3/1/0.3 µg/mL and the fragments were diluted to corresponding antigen-binding molarities and analyzed as above.

mAb 3A2 was also used as quantification control. For this purpose, purified mAb was spiked into normal human serum at concentrations of 10, 2.5, or 0.5 µg/mL; normal serum was left unspiked as the negative control. The preparations were aliquoted and frozen, and freshly thawed aliquots were used to monitor run-to-run and day-to-day assay performance.

### Samples

Positive and negative human serum and plasma samples were sourced via BioIVT, iSpecimen, StemExpress and Emory University. Time to collection from symptom onset was available for 129 of 186 positive serum controls and varied from 6 to 71 days, with 10% and 90% percentiles at 15 and 53 days, respectively. Additionally, residual serum samples collected for hantavirus and Zika virus serology prior to emergence of SARS-CoV-2 were tested as negative controls (as part of the training set and the validation set, respectively). The hantavirus samples were inactivated by 5 × 10^6^ RAD gamma irradiation from a ^60^Co source. All samples were heated to 56°C for 10 min before use. The serum samples from commercial sources were analyzed in a blinded fashion. Approximately 6% of initially analyzed samples were excluded from final analysis after identifying multiple samples from the same donor (pre-outbreak residual samples) or due to inconclusive information regarding previous SARS-CoV-2 nucleic acid testing results (commercially sourced samples after de-blinding). Residual specimen materials were used for diagnostics development under a protocol that was reviewed and approved by the CDC Institutional Review Board^§^.

### Purification of IgG and IgM from human serum

Human serum samples from SARS-CoV-2 positive donors or control donors (N = 14 each) were pooled, and IgG and IgM antibodies were purified according to manufacturer’s recommendations using CaptureSelect CH1-Xl and Poros CaptureSelect IgM Affinity matrices (Thermo Fisher Scientific). The samples were concentrated and the buffer was changed to PBS using 10.000 Da spin columns (Amicon). The purified fractions were analyzed by SDS-PAGE both under non-heated, non-reduced conditions and under heated, reduced conditions.

### Statistics

Statistical analyses were performed using GraphPad Prism software version 7.04 except for the kappa statistic, which was determined using the online version of GraphPad at https://www.graphpad.com/quickcalcs/kappa1.cfm (confidence interval calculated with equations by Fleiss).

## Data Availability

No external data links

## Acknowledgements

We thank Tatyana Klimova for assistance in editing this manuscript; John Klena, Inna Krapiunaya and Jarad Schiffer for assistance with sample access and the Laboratory Task Force of the CDC COVID-19 response for their critical project review and resource support. This research was made possible using samples obtained from the CDC Biorepository. The findings and conclusions in this report are those of the authors and do not necessarily represent the official position of the Centers for Disease Control and Prevention.

## Footnotes

§ See 45 C.F.R. part 46; 21 C.F.R. part 56.

## Author contributions

MHK and EB designed and conducted experiments and contributed to the manuscript; PC and HMJ conducted experiments; APC, JL, AC, XT, REW, KBM, MGF and JMG developed monoclonal antibodies and mouse sera; MP, AJS, PSS, JMC and SMO acquired samples and compiled associated data; TDF reviewed statistics; JMM and CFS managed the project and contributed to the manuscript.

## Materials availability

unique research materials will be shared upon completion of appropriate material transfer agreements.

## Supplementary Materials

**Supplementary Fig. 1:**
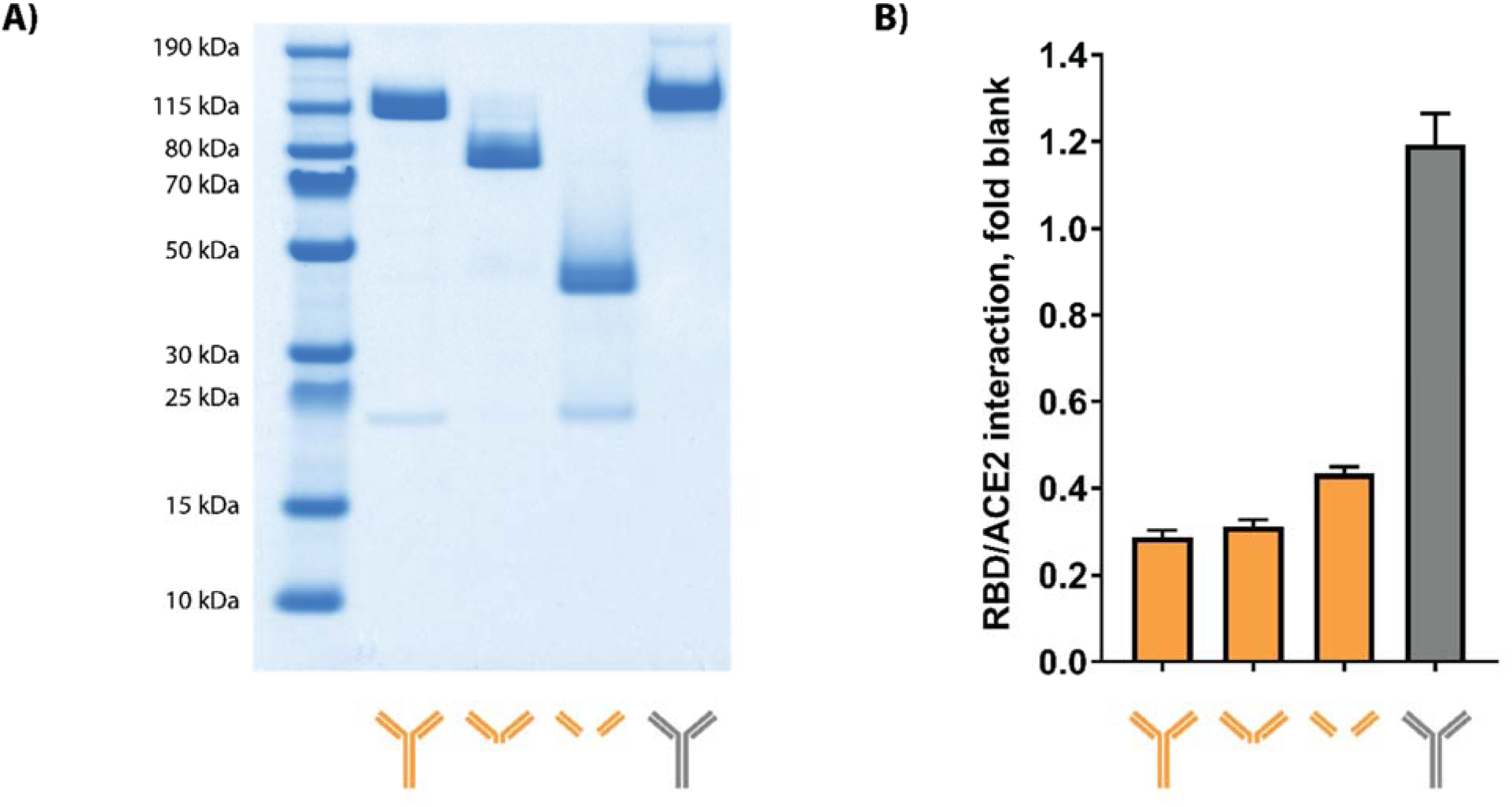
mAbs, F(ab’)2 and Fab fragments analyzed by SDS-PAGE; RBD/ACE2 inhibition assay. A) SARS-CoV-2 RBD-specific mAb 3A2 (orange) was analyzed with non-reducing, non-heated SDS-PAGE, both intact and after cleavege to F(ab’)2 and Fab fragments as described in Materials and Methods. A non-targeting isotype control mAb (gray) was analyzed in parallel. B) The ability of 3A2 mAb, 3A2 F(ab’)2, 3A2 Fab, and isotype control mAb to inhibit the interaction between SARS-CoV-2 RBD and its receptor ACE2 was quantified using the assay described in Materials and Methods.

**Supplementary Fig. 2:**
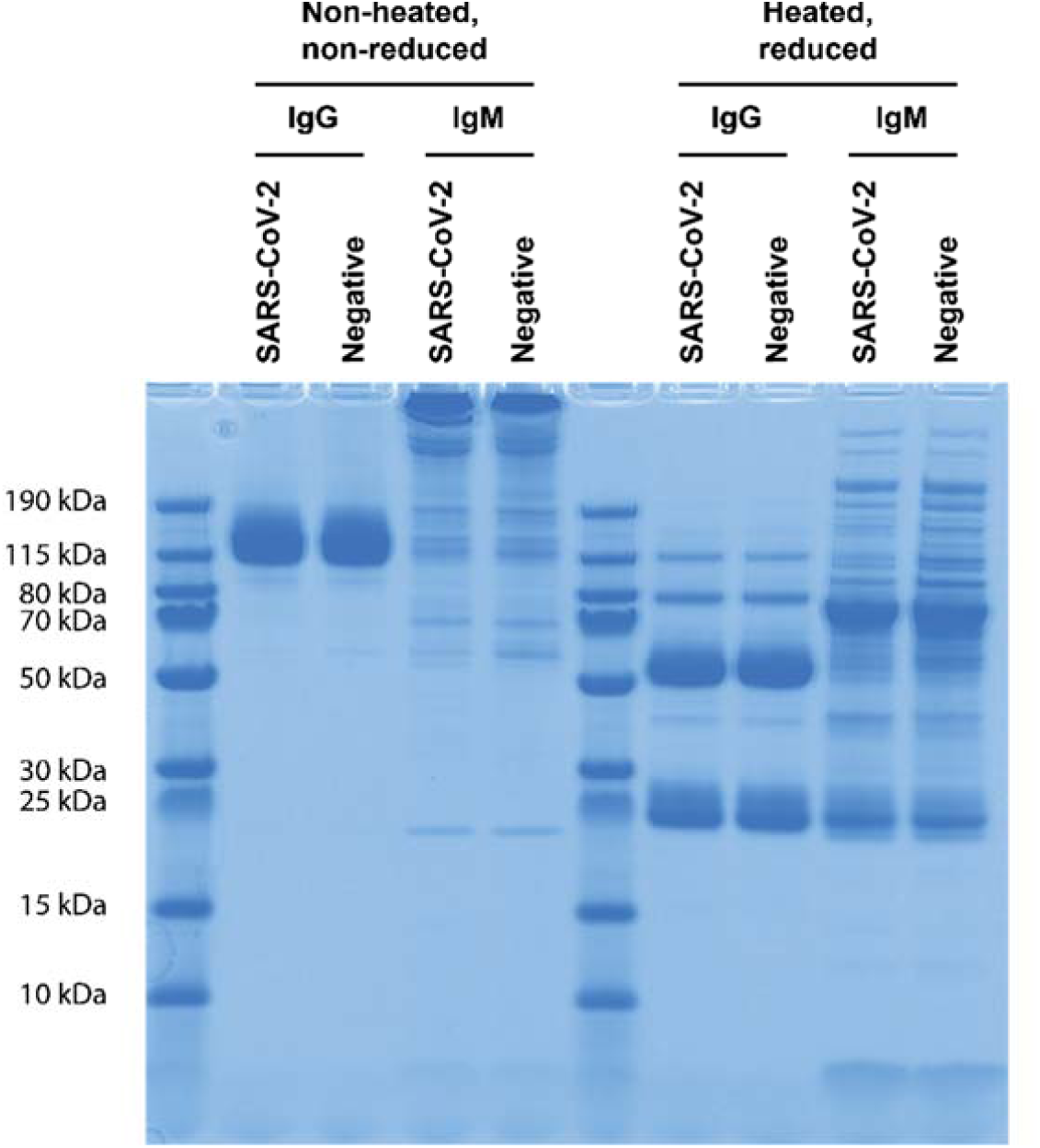
IgG and IgM antibodies purified from pooled sera of convalescent Covid-19 patients or from a control serum pool. The purification products (10 pg/lane) were separated by SDS-PAGE on a 4-12% Bis-Tris gel with MES running buffer and stained with InstantBlue Coomassie stain.

**Supplementary Fig. 3:**
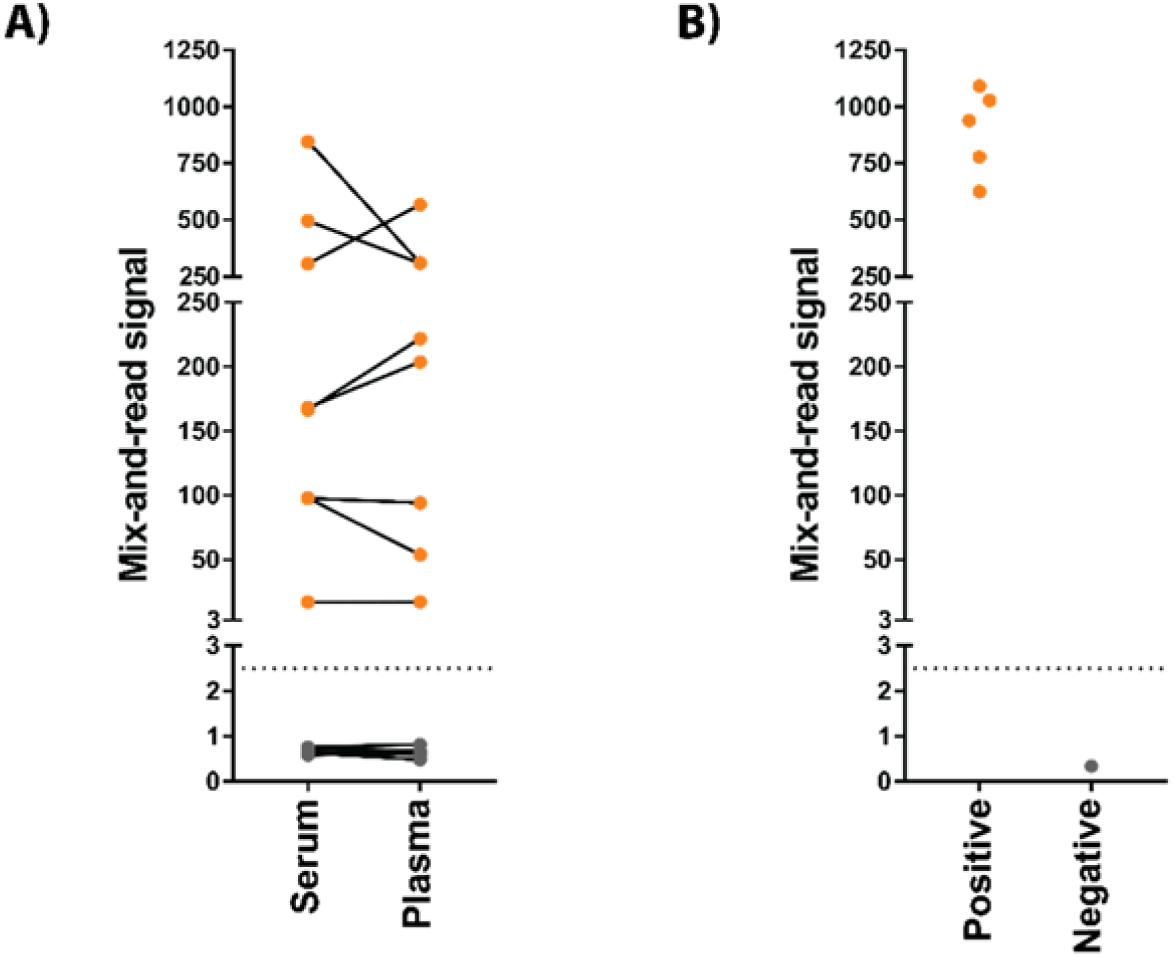
Comparing serum and plasma as sample matrices; analyzing mouse serum. A) Paired human serum and EDTA plasma samples from 8 SARS-CoV-2-positive and 8 negative individuals were obtained from commercial sources and tested in parallel by the mix-and-read assay. Orange: samples from individuals with history of PCR-confirmed SARS-CoV-2 infection. Gray: negative control samples. B) Analyzing mouse serum. Five serum samples from mice that were repeatedly immunized in order to generate antibodies against SARS-CoV-2 spike domains were analyzed as positive controls, and negative control serum was from an untreated mouse.

**Supplementary Fig. 4:**
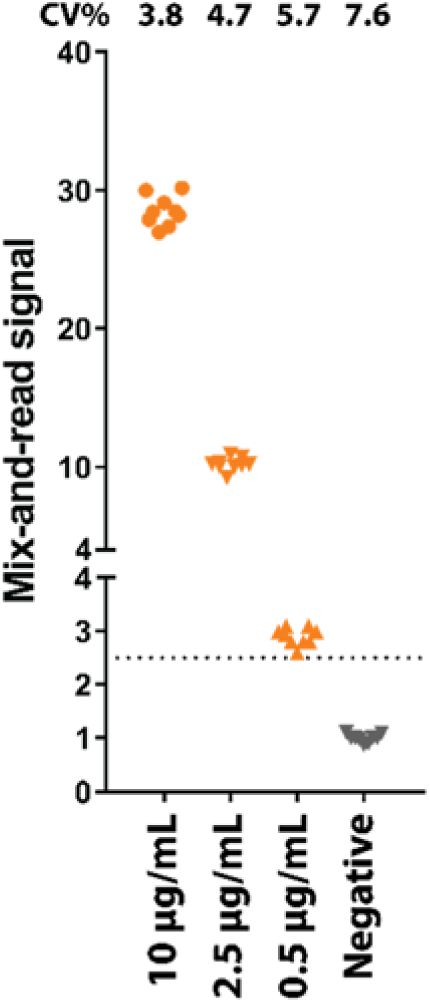
Run-to-run reproducibility of quantification controls. 3A2 mAb was spiked into normal human serum in 3 different concentrations, or serum was left without mAb for a negative control. Aliquots were stored frozen and run together with samples of interest to monitor day-to-day and run-to-run performance of the assay. Data from 9 independent runs presented. CV; coefficient of variation.

**Supplementary Fig. 5.**
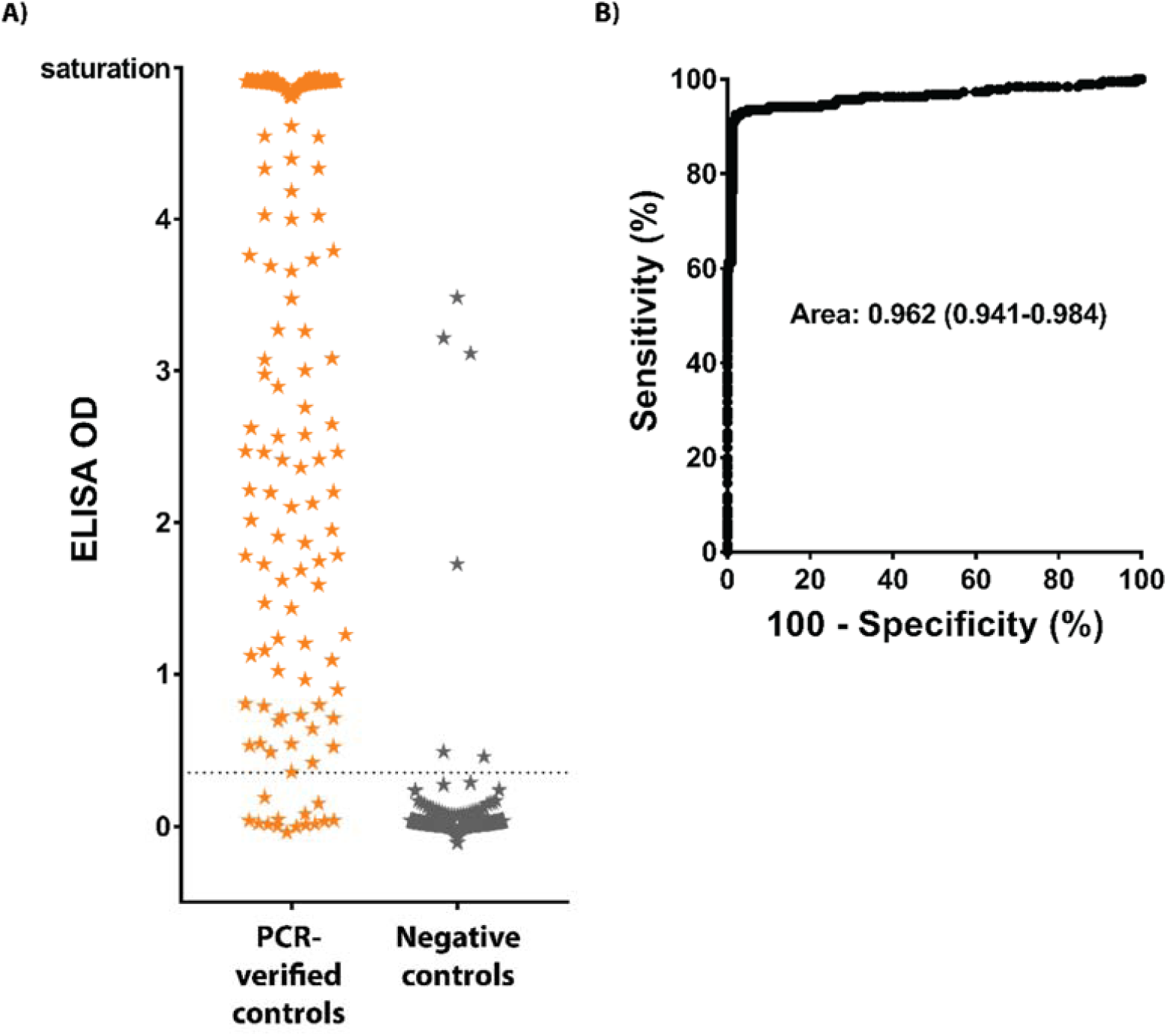
Compiled RBD ELISA results. A) OD distribution of the training set and validation set samples using 1:100 screening dilution (N = 186 samples positive for SARS-CoV-2 by nucleic acid testing and N = 300 negative controls). Values were corrected by reducing signal from wells with no antigen. B) ROC analysis. Cut-off value of 0.35 enables 92% sensitivity and 98% specificity.

